# Topical anticholinergic medications for primary hyperhidrosis: A protocol for a systematic review and meta-analysis

**DOI:** 10.1101/2022.03.05.22271966

**Authors:** Yang Wu, Wenli Gao, Jiao Yan, Min Yang, Dongyang Gao

**Affiliations:** Department of Integrated Chinese and Western Medicine, The General Hospital of Western Theater Command, Chengdu, Sichuan, P.R. China; Tianjin University of Traditional Chinese Medicine, Tianjin, China; School of Life Sciences, Beijing University of Chinese Medicine, Beijing, China

**Author notes:** Correspondence (M.Y.); (D.G.). **Yang Wu (first author):**. **Wenli Gao(first author):**. **Jiao Yan:**. These authors have contributed equally to this work and share first authorship (Yang Wu, Wenli Gao).

**Keywords:** Primary hyperhidrosis, anticholinergic drugs, topical, systematic review

## Abstract

**Background:** Primary hyperhidrosis (PHH) is a chronic condition characterized by excessive sweating. Several topical anticholinergic agents have been developed, but the evidence for the efficacy and tolerability of changing medication pathways of anticholinergics for PHH is limited. We will review the available literature to derive information regarding the efficacy and safety of different medicines used for the topical treatment of patients with hyperhidrosis.

**Methods:** PubMed, Cochrane Library, Embase, Web of Science, and Cochrane Central Register of Controlled Trials will be searched from inception to May 2023 for studies that may be eligible for inclusion. Randomized controlled trials (RCTs) of PHH treated with topical anticholinergic drugs will be included in the analysis. The primary outcomes will include the severity of hyperhidrosis measured quantitatively, the Hyperhidrosis Disease Severity Scale (HDSS) score, or the proportion of subjects with a minimum 2-grade improvement from baseline in HDSS, the Hyperhidrosis Disease Severity Measure-Axillary (HDSM-Ax) score, and Dermatology Life Quality Index (DLQI). The secondary outcomes will focus on safety and tolerability. Study selection, data extraction, and assessment of the risk of bias will be independently performed by two investigators. Data synthesis will be performed using Review Manager 5.4 software.

**Results:** The results will be published as a peer-reviewed article.

**Conclusions:** This review will systematically evaluate the existing evidence on the efficacy and safety of topical anticholinergic agents in the treatment of PHH. These conclusions will provide supporting evidence for the current guidelines and more reliable evidence-based medicine references for clinical practitioners and researchers.

**PROSPERO registration number:** CRD42022312316

## Introduction

Hyperhidrosis (HH) is a chronic disease in which the degree of sweating exceeds the physiological requirements for maintaining internal temperature regulation [1, 2]. HH is characterized by excessive local or systemic sweating due to exocrine sweat gland hyperfunction [3, 4], commonly found in areas such as axillae, palms, soles, and face [5, 6]. Hyperhidrosis is divided into primary hyperhidrosis (PHH) and secondary hyperhidrosis, based on whether there is an underlying disease or drug use [2, 7]. The prevalence of HH ranges from 1% to 4.8% [8, 9], and is seen in both in children and adults. The average age of onset is 14-25 years [10]. The discomfort associated with severe hyperhidrosis can interfere with daily life and even lead to social and work isolation [11-13], with significant negative effects on the patient’s daily activities, mental health, and professional behavior [14, 15].

The pathophysiology, diagnosis, and treatment of PHH have attracted considerable attention. Perspiration is produced by the secretion of sweat glands innervated by the cholinergic nerve fibers of the sympathetic nervous system [16]. Studies suggest that hyperactivity of cholinergic nerve fibers in sweat glands may lead to PHH [17]. In addition, an excessive number or abnormal distribution of sweat glands may also be the cause of the disease [18]. The anterior cingulate cortex, which controls responses to emotional stimuli, also exerts a significant influence [19].

Management of PHH includes systemic, local, psychological, and surgical treatments. [16] Systemic therapies include oral anticholinergic drugs [20] such as oxybutynin [21]. Topical treatment includes the use of alumina, which chemically blocks sweat ducts to reduce sweating [22], as well as topical anticholinergic agents, such as glycopyrronium bromide. Ion import at specific sites is usually the treatment of choice for HH of the palms and soles, but long-term maintenance treatment is required [23]. Botulinum toxin type A (Botox, approved by the Food and Drug Administration (FDA) in 2004 [6] and subsequently by several countries [24] is the first- or second-line treatment option for axillary and plantar hyperhidrosis [25]. This toxin blocks cholinergic stimulation of sweat glands. Psychotherapy is beneficial in some cases [26, 27]. Surgery, including local surgery and endoscopic thoracic sympathetic nerve resection, can be considered when conservative treatment fails [7, 28].

However, these treatments are associated with side effects and complications. Adverse events associated with oral anticholinergics, such as dry mouth, urinary retention, constipation, and headache [29], often lead to interruption of drug use and even increase the risk of cognitive impairment [30]. Studies have shown that its efficacy decreases with long-term treatment [31]. Other adverse effects include skin irritation, pain, and burns caused by ion import [32], repeated injections of Botox [16], and compensatory sweating after sympathetic nerve resection [33, 34].

Therefore, improving the tolerability of preparations has become the focus of research, especially in the development of preparations for external use [35]. Several topical anticholinergic drugs have been approved in recent years. Glycopyrronium tosylate (Qbrexza™, a pre-wetting cloth containing 2.4% glycopyrronium solution) reduces sweating by blocking the activation of acetylcholine receptors in the peripheral sweat glands [15] and was approved by the FDA in 2018 [36]. Topical sofpironium bromide(ECCLOCK®, 5%) gel reduces sweat secretion by inhibiting local sweat gland M3 muscarinic receptor [37], it was approved in Japan in September 2020. In addition, topical oxybutynin chloride cream or gel formulations have shown good tolerability, and promise to be of benefit in the future [38]. Topical preparation of glycopyrronium bromide has aroused interest as a treatment option for PHH [39], and 1% glycopyrronium bromide cream has completed the phase IIIa study [40]. The dermal preparation of umeclidinium ammonium bromide is also under way [41].

Previous meta-analyses have assessed the efficacy and safety of oral anticholinergic agents (oxybutynin, methane bromide, glycopyrrolate, etc.) for the treatment of PHH and have shown that improved outcomes are associated with a large number of adverse events [5]. There are currently several topical anticholinergic agents on the market, and the evidence for the efficacy and tolerability of a change in the route of administration in the treatment of PHH has not been thoroughly evaluated. Therefore, systematic reviews and meta-analyses are necessary to provide supporting evidence for the current guidelines and more reliable evidence-based medicine references for clinical practitioners and researchers.

## Materials and Methods

### Study registration

This meta-analysis protocol follows the Preferred Reporting Items for Systematic Review and Meta-Analysis Protocols (PRISMA-P) [42] and the Manual for Systematic Evaluation of Cochrane Interventions. The PRISMA-P checklist is in S1 Appendix. The protocol has been registered with the International Prospective Register of Systematic Reviews (PROSPERO) (CRD42022312316). The investigators will receive consistent training to ensure that they are familiar with the background, purpose, and methodology of the system evaluation.

### Eligibility criteria

#### Type of studies

Randomized controlled trials (RCTs) of PHH treated with topical anticholinergic drugs will be included. Completed trials published on a clinical trial registration platform will also be included. Quasi-randomized trials will be excluded.

#### Types of participants

Participants with a diagnosis of primary hyperhidrosis based on focal, visible, excessive sweating for at least 6 months without apparent cause, and meeting two or more of the following criteria at the same time: (1) onset age of 25 years or younger, (2) bilateral and relatively symmetric distribution, (3) at least one episode weekly, (4) cessation of sweating during sleep, (5) impaired daily activities, and (6) family history of primary hyperhidrosis will be included. Participants will have to have a hyperhidrosis disease severity scale (HDSS) score of at least three during the baseline evaluation.

Patients with secondary hyperhidrosis will be excluded from this study. Patients will be excluded if they have another concurrent cutaneous or subcutaneous disease or if they have received any treatment or undergone any procedure that could interfere with drug activity.

#### Type of interventions and controls

RCTs comparing topical anticholinergics such as glycopyrronium tosylate, sofpironium bromide, umeclidinium, or oxybutinin with placebo, and RCTs comparing two types of aforementioned topical anticholinergics will be included. Trials comparing the same type of anticholinergic drugs at different concentrations or treatment durations will be considered as eligible. Trials involving combination therapy (e.g., a combination of topical and oral anticholinergic drugs) will be excluded.

#### Type of outcome measures

Primary outcomes: (1) Severity of hyperhidrosis measured quantitatively (e.g., gravimetry, evaporimeter, and minor starch-iodine test). (2) Hyperhidrosis Disease Severity Scale (HDSS) score or the proportion of subjects with a minimum 2-grade improvement from baseline in the HDSS. (3) Hyperhidrosis Disease Severity Measure-Axillary (HDSM-Ax) score. (4) Dermatology Life Quality Index (DLQI). Secondary outcomes: Safety and tolerability will be evaluated using the following outcomes: (1) total number of patients who experienced treatment-emergent adverse events (TEAEs) such as blurred vision or mydriasis, dry mouth, urinary retention, constipation, anhidrosis, dizziness, headache, nasopharyngitis, etc.; (2) total number of patients who experienced local skin reactions (LSRs), including burning, stinging, pruritus, edema, erythema, dryness, and scaling; and (3) total number of patients who experienced serious adverse events(according to the classification employed by the US FDA).

#### Data sources and search strategies

PubMed, Cochrane Library, Embase, Web of Science, and Cochrane Central Register of Controlled Trials will be searched from inception to May 2023 for studies that may be eligible for inclusion. In addition, we will identify other studies that meet the inclusion criteria from the list of references included in the study. Clinical trial registries, such as clinicaltrials.gov and WHO ICTRP, will also be searched for ongoing or unpublished clinical trials.

Two investigators (YW and WG) independently searched the literature using a combination of medical subject heading (MeSH) terms and free-text terms, and designed a search formula for each electronic database, including “hyperhidrosis,” “anticholinergic drugs,” and “randomized controlled trials”. There were no language restrictions in the search process. PubMed’s search strategy is shown in S2 Appendix and will be modified to meet the needs of other databases.

#### Studies selection

The literature will be screened independently by two investigators (YW and WG) according to pre-established inclusion and exclusion criteria. The search results will be imported into Endnote X9.0, and duplicated data will be deleted. The titles and abstracts of the studies were read independently to screen out irrelevant studies and records. The full paper will then be read and non-randomized controlled trials, unqualified controls, and duplicate studies will be excluded. Any discrepancies in the screening process will be resolved through discussion with a third investigator (DG). The details of study selection are summarized in a flow chart (Figure 1).

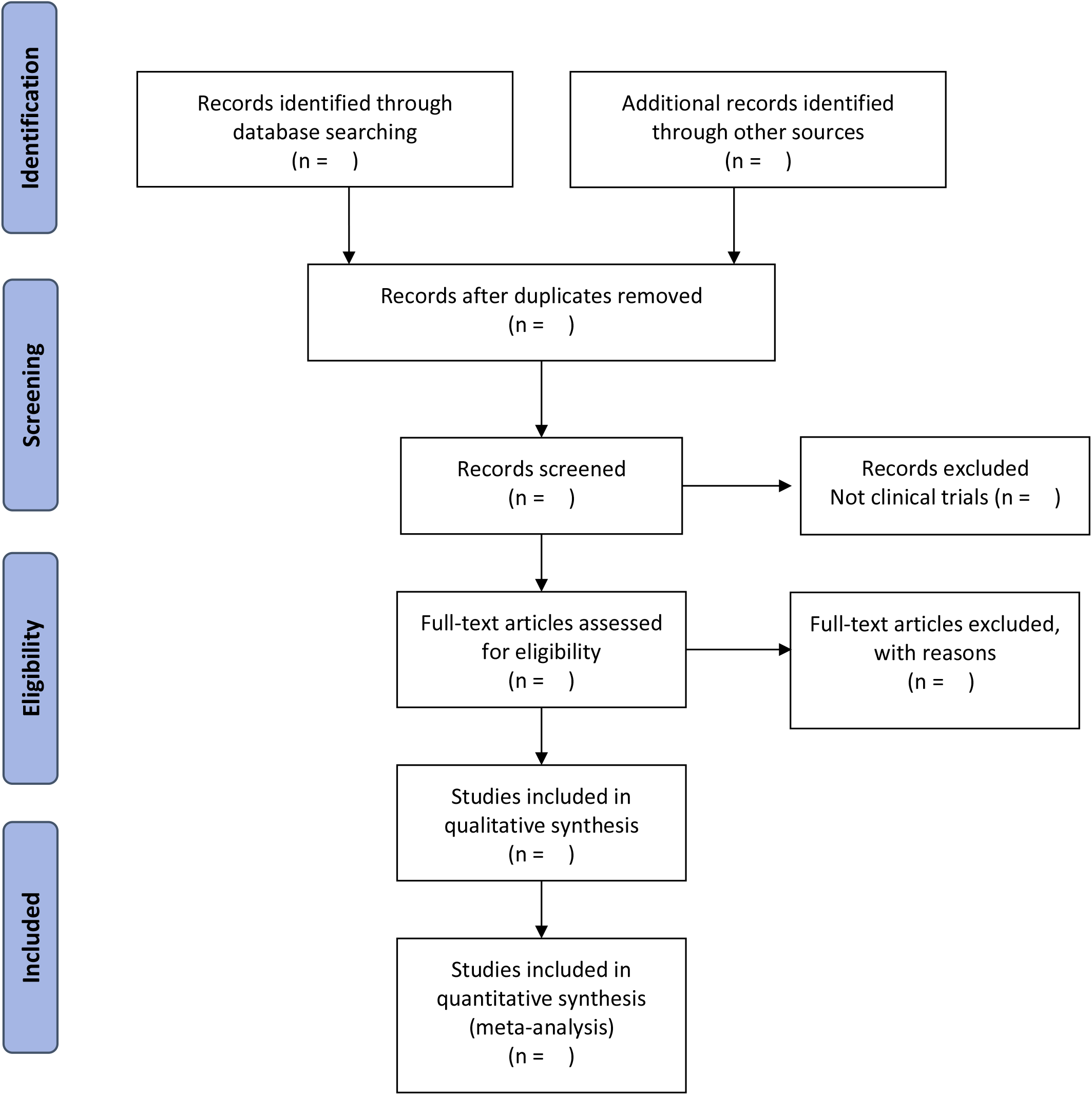

#### Data extraction and management

Two investigators (WG and JY) independently extracted the data using standard Excel spreadsheets, including the first author, year of publication, country, research type, research design, baseline information for each group, intervention, treatment time, outcome indicators, and adverse reactions. The extracted data will be crosschecked. If there is any discrepancy, the original article will be checked and the discrepancy resolved by consensus. When data are not reported in a usable format and are difficult to reliably extract, the authors will be contacted by email for information twice over four weeks. If some data are still not available from authors, they can be acquired based on available data referring to advanced methods of data extraction, such as intercepting data from a statistical graph and data conversion. [43]

#### Assessment of risk of bias

The Cochrane Risk Assessment of Bias Tool [44] will be applied by two investigators (YW and WG) to independently assess the risk of bias in the included RCTs. The assessment includes random sequence generation, allocation concealment, blinding of participants and personnel, blinding of outcome assessment, incomplete outcome data, selective reporting, and other biases. The criteria used to assess the risk of bias are “low risk,” “high risk” and “unclear [44]”. Disagreements regarding the risk of bias will be resolved by consultation with a third investigator (DG). A bias risk assessment chart will be generated to present the assessment results and details.

#### Statistical analysis

Review Manager 5.4 software [42] (Copenhagen: The Nordic Cochrane Centre, The Cochrane Collaboration) will be used for data synthesis and statistical analysis. For dichotomous variables, relative risk (RR), risk difference (RD), 95% confidence interval (CI), and P-values will be calculated. For continuous variables, the mean difference (MD) or standard mean difference (SMD), 95% confidence interval (CI), and P-values will be calculated using the random-effects model. Simultaneously, the Q test and I2 index will be used to assess statistical heterogeneity. When heterogeneity is significant (I2>50%), we will first explore the possible sources of heterogeneity. A sensitivity analysis will be performed to examine the sources of heterogeneity. When there is clinical or methodological heterogeneity, subgroup analysis or meta-regression(with Stata V.16.0 software) will be performed. [42] When there is statistical heterogeneity, a random-effects model will be used. Finally, if considerable heterogeneity cannot be resolved, a systematic review of descriptive analyses will be performed instead of a meta-analysis.

#### Dealing with missing data

If there is missing data from the included RCT studies, the original articles, supplemental materials, and even other articles published based on the same trial will be scrutinized again for omissions. Otherwise, the investigators will attempt to contact the corresponding author of the study by email twice over 4 weeks. If complete data are not available, the study will not be included. The potential impact of missing data will be assessed using a sensitivity analysis, as described in the results.

#### Subgroup analysis and sensitivity analysis

Subgroup analysis based on different comparisons and different skin sites (forehead, axillae, palms, etc.) where the drugs are applied will be conducted. If there is significant heterogeneity, the trials will be further classified to perform a subgroup analysis after exploration. A sensitivity analysis will be performed to evaluate the robustness of the results and to explore the source of heterogeneity. In addition, given the different results between the same risk of bias domain of studies, studies can be divided into different subgroups based on “low risk”, “high risk of bias”, and “not clear” to compare the difference in the estimate of effect and analyze the contribution of subgroups to the pooled effect size, which is conducive to grade the quality of evidence.

#### Publication bias

When more than 10 studies met the inclusion criteria, we will evaluate the potential publication bias by drawing funnel plots (with Review Manager 5.4 software) and Egger tests (with Stata V.16.0 software).

#### Grading the quality of evidence

We will evaluate the evidence quality of the included studies according to the Grading of Recommendations Assessment, Development, and Evaluation (GRADE) approach [45] The evidence quality GRADE is divided into four grades: high, moderate, low, and very low.

## Results

The results of this systematic review will be published as a peer-reviewed article.

## Discussion

Although there are many therapeutic options for PHH, treatment plans should be individualized according to the actual situation of patients[35]. Clinical guidelines for PHH no longer recommend systemic therapy because of unacceptable adverse reactions to oral anticholinergic drugs in recent years[46]. Topical anticholinergic agents optimize existing therapies and are likely to replace systemic oral administration[17].

This review will systematically evaluate the existing evidence on the efficacy and safety of topical anticholinergic agents in the treatment of PHH. These conclusions will help clinicians or practitioners to develop individual and more optimal treatment options for patients with PHH and provide supporting evidence for health policymakers and the current guidelines.

## Conclusions

This systematic review aims to provide comprehensive evidence on the efficacy and safety of topical anticholinergic agents for primary hyperhidrosis. The findings of this review may help point to better forms of treatment based on evidence-based medicine.

## Supporting information

S1 Appendix. PRISMA-P-checklist

S2 Appendix. The search strategy for PubMed

## Data Availability

All data produced in the present study are available upon reasonable request to the authors

## Author Contributions

Conceptualization, D.G. and Y.W.; methodology, W.G.; software, D.G.; validation, W.G., M.Y. and Y.W.; formal analysis, D.G.; investigation, Y.W.; resources, Y.W.; data curation, W.G. and J.Y.; writing—original draft preparation, Y.W. and W.G.; writing—review and editing, D.G.; visualization, D.G.; supervision, M.Y.; project administration, M.Y.; funding acquisition, M.Y. All authors have read and agreed to the published version of the manuscript.”

## Supporting information

## S1 Appendix.

The PRISMA-P checklist.

## S2 Appendix.

The search strategy for PubMed.

## References

1. Caserta F, Lenn J, Hofland H. Variables Affecting Delivery of Glycopyrronium Tosylate Through Human Skin In Vitro. J Drugs Dermatol. 2020;19(11):1080–5. Epub 2020/11/17. doi: 10.36849/jdd.2020.5062. PubMed PMID: 33196749.

2. Liu V, Farshchian M, Potts GA. Management of Primary Focal Hyperhidrosis: An Algorithmic Approach. J Drugs Dermatol. 2021;20(5):523–8. Epub 2021/05/04. doi: 10.36849/jdd.5774. PubMed PMID: 33938689.

3. Wolosker N, Kauffman P, de Campos JRM, Faustino CB, da Silva MFA, Teivelis MP, et al. Long-term results of the treatment of primary hyperhidrosis with oxybutynin: follow-up of 1,658 cases. Int J Dermatol. 2020;59(6):709–15. Epub 2020/04/18. doi: 10.1111/ijd.14872. PubMed PMID: 32301117.

4. Basedow S, Kruse R, Bruch-Gerharz D. [Hyperhidrosis of childhood and adolescence: clinical aspects and therapeutic options]. Hautarzt. 2011;62(12):928–34. Epub 2011/10/12. doi: 10.1007/s00105-011-2225-7. PubMed PMID: 21987173.

5. Cruddas L, Baker DM. Treatment of primary hyperhidrosis with oral anticholinergic medications: a systematic review. J Eur Acad Dermatol Venereol. 2017;31(6):952–63. Epub 2016/12/16. doi: 10.1111/jdv.14081. PubMed PMID: 27976476.

6. Cohen JL, Cohen G, Solish N, Murray CA. Diagnosis, impact, and management of focal hyperhidrosis: treatment review including botulinum toxin therapy. Facial Plast Surg Clin North Am. 2007;15(1):17-30, v-vi. Epub 2007/02/24. doi: 10.1016/j.fsc.2006.10.002. PubMed PMID: 17317552.

7. McConaghy JR, Fosselman D. Hyperhidrosis: Management Options. Am Fam Physician. 2018;97(11):729–34. Epub 2018/09/15. PubMed PMID: 30215934.

8. Delort S, Marchi E, Corrêa MA. Oxybutynin as an alternative treatment for hyperhidrosis. An Bras Dermatol. 2017;92(2):217–20. Epub 2017/05/26. doi: 10.1590/abd1806-4841.201755126. PubMed PMID: 28538882; PubMed Central PMCID: PMCPMC5429108.

9. Ram R, Lowe NJ, Yamauchi PS. Current and emerging therapeutic modalities for hyperhidrosis, part 1: conservative and noninvasive treatments. Cutis. 2007;79(3):211–7. Epub 2007/08/07. PubMed PMID: 17674587.

10. Kumar MG, Foreman RS, Berk DR, Bayliss SJ. Oral glycopyrrolate for refractory pediatric and adolescent hyperhidrosis. Pediatr Dermatol. 2014;31(1):e28–30. Epub 2013/11/26. doi: 10.1111/pde.12236. PubMed PMID: 24266878.

11. Callejas MA, Grimalt R, Cladellas E. [Hyperhydrosis update]. Actas Dermosifiliogr. 2010;101(2):110–8. Epub 2010/03/13. doi: 10.1016/j.ad.2009.09.004. PubMed PMID: 20223153.

12. Bajaj V, Langtry JA. Use of oral glycopyrronium bromide in hyperhidrosis. Br J Dermatol. 2007;157(1):118–21. Epub 2007/04/27. doi: 10.1111/j.1365-2133.2007.07884.x. PubMed PMID: 17459043.

13. Nyamekye IK. Current therapeutic options for treating primary hyperhidrosis. Eur J Vasc Endovasc Surg. 2004;27(6):571–6. Epub 2004/05/04. doi: 10.1016/j.ejvs.2004.01.023. PubMed PMID: 15121105.

14. Rzany B, Müller C, Hund M. [Focal hyperhidrosis.Quality of life, socioeconomic importance and use of internal medicinal therapy]. Hautarzt. 2012;63(6):456–61. Epub 2012/06/20. doi: 10.1007/s00105-012-2331-1. PubMed PMID: 22710840.

15. Nwannunu CE, Limmer AL, Coleman K, Shah R, Patel RR, Mui UN, et al. Glycopyrronium Tosylate (Qbrexza) for Hyperhidrosis. Skin Therapy Lett. 2019;24(2):1–3. Epub 2019/04/11. PubMed PMID: 30970203.

16. Connolly M, de Berker D. Management of primary hyperhidrosis: a summary of the different treatment modalities. Am J Clin Dermatol. 2003;4(10):681–97. Epub 2003/09/26. doi: 10.2165/00128071-200304100-00003. PubMed PMID: 14507230.

17. Wechter T, Feldman SR, Taylor SL. The Treatment of Primary Focal Hyperhidrosis. Skin Therapy Lett. 2019;24(1):1–7. Epub 2019/03/01. PubMed PMID: 30817880.

18. Sammons JE, Khachemoune A. Axillary hyperhidrosis: a focused review. J Dermatolog Treat. 2017;28(7):582–90. Epub 2017/03/21. doi: 10.1080/09546634.2017.1309347. PubMed PMID: 28318360.

19. Eisenach JH, Atkinson JL, Fealey RD. Hyperhidrosis: evolving therapies for a well-established phenomenon. Mayo Clin Proc. 2005;80(5):657–66. Epub 2005/05/13. doi: 10.4065/80.5.657. PubMed PMID: 15887434.

20. Baumgartner FJ, Bertin S, Konecny J. Superiority of thoracoscopic sympathectomy over medical management for the palmoplantar subset of severe hyperhidrosis. Ann Vasc Surg. 2009;23(1):1–7. Epub 2008/07/16. doi: 10.1016/j.avsg.2008.04.014. PubMed PMID: 18619780.

21. Nicholas R, Quddus A, Baker DM. Treatment of Primary Craniofacial Hyperhidrosis: A Systematic Review. Am J Clin Dermatol. 2015;16(5):361–70. Epub 2015/06/10. doi: 10.1007/s40257-015-0136-6. PubMed PMID: 26055729.

22. Pariser DM, Ballard A. Topical therapies in hyperhidrosis care. Dermatol Clin. 2014;32(4):485–90. Epub 2014/08/26. doi: 10.1016/j.det.2014.06.008. PubMed PMID: 25152341.

23. Kim WO, Kil HK, Yoon DM, Cho MJ. Treatment of compensatory gustatory hyperhidrosis with topical glycopyrrolate. Yonsei Med J. 2003;44(4):579–82. Epub 2003/09/02. doi: 10.3349/ymj.2003.44.4.579. PubMed PMID: 12950111.

24. Murray CA, Cohen JL, Solish N. Treatment of focal hyperhidrosis. J Cutan Med Surg. 2007;11(2):67–77. Epub 2007/03/22. doi: 10.2310/7750.2007.00012. PubMed PMID: 17374317.

25. Reisfeld R, Berliner KI. Evidence-based review of the nonsurgical management of hyperhidrosis. Thorac Surg Clin. 2008;18(2):157–66. Epub 2008/06/19. doi: 10.1016/j.thorsurg.2008.01.004. PubMed PMID: 18557589.

26. Glaser DA, Hebert AA, Pariser DM, Solish N. Primary focal hyperhidrosis: scope of the problem. Cutis. 2007;79(5 Suppl):5–17. Epub 2007/06/29. PubMed PMID: 17596096.

27. Maillard H, Lecouflet M. [Management of hyperhidrosis]. Ann Dermatol Venereol. 2015;142(4):252–61. Epub 2015/02/24. doi: 10.1016/j.annder.2014.11.005. PubMed PMID: 25697587.

28. Nawrocki S, Cha J. The etiology, diagnosis, and management of hyperhidrosis: A comprehensive review: Therapeutic options. J Am Acad Dermatol. 2019;81(3):669–80. Epub 2019/02/03. doi: 10.1016/j.jaad.2018.11.066. PubMed PMID: 30710603.

29. Campanati A, Gregoriou S, Consales V, Rizzetto G, Bobyr I, Diotallevi F, et al. Combined treatment of palmar hyperhidrosis with botulinum toxin type A and oxybutynin chloride: Results of a clinical, multicenter, prospective study. Dermatologic Therapy. 2020;33(6):e14039. doi: 10.1111/dth.14039. PubMed PMID: 32691938.

30. Gregoriou S, Kontochristopoulos G, Tiligada E. Pharmacokinetic profile data of glycopyrronium bromide 1% cream beyond 2 weeks are important. The British Journal of Dermatology. 2021;185(2):467–8. doi: 10.1111/bjd.20088. PubMed PMID: 33792896.

31. Del Boz Gonzalez J, Rodríguez Barón D, Millán-Cayetano JF, de Troya Martin M. Tolerance of oral oxybutynin in the treatment of hyperhidrosis. Dermatol Ther. 2020;33(6):e14197. Epub 2020/08/15. doi: 10.1111/dth.14197. PubMed PMID: 32794595.

32. Shimizu H, Tamada Y, Shimizu J, Ohshima Y, Matsumoto Y, Sugenoya J. Effectiveness of iontophoresis with alternating current (AC) in the treatment of patients with palmoplantar hyperhidrosis. J Dermatol. 2003;30(6):444–9. Epub 2003/06/18. doi: 10.1111/j.1346-8138.2003.tb00414.x. PubMed PMID: 12810991.

33. Cladellas E, Callejas MA, Grimalt R. A medical alternative to the treatment of compensatory sweating. Dermatol Ther. 2008;21(5):406–8. Epub 2008/10/11. doi: 10.1111/j.1529-8019.2008.00222.x. PubMed PMID: 18844718.

34. Costa Ada S, Jr., Leão LE, Succi JE, Perfeito JA, Filho Castelo A, Rymkiewicz E, et al. Randomized trial - oxybutynin for treatment of persistent plantar hyperhidrosis in women after sympathectomy. Clinics (Sao Paulo). 2014;69(2):101–5. Epub 2014/02/13. doi: 10.6061/clinics/2014(02)05. PubMed PMID: 24519200; PubMed Central PMCID: PMCPMC3912338.

35. Brown AL, Gordon J, Hill S. Hyperhidrosis: review of recent advances and new therapeutic options for primary hyperhidrosis. Curr Opin Pediatr. 2014;26(4):460–5. Epub 2014/06/07. doi: 10.1097/mop.0000000000000108. PubMed PMID: 24905102.

36. Glaser DA, Hebert AA, Nast A, Werschler WP, Green L, Mamelok RD, et al. A 44-Week Open-Label Study Evaluating Safety and Efficacy of Topical Glycopyrronium Tosylate in Patients with Primary Axillary Hyperhidrosis. Am J Clin Dermatol. 2019;20(4):593–604. Epub 2019/05/22. doi: 10.1007/s40257-019-00446-6. PubMed PMID: 31111409; PubMed Central PMCID: PMCPMC6687675 Biotech, Inc., Dermira, Inc., Evolus, and Sienna Biopharmaceuticals, Inc. and a consultant for Dermira, Inc. William P. Werschler is a consultant and investigator for Dermira, Inc. Adelaide A. Herbert is a consultant for Dermira, Inc. and an employee of the UTHealth McGovern Medical School, which received compensation from Dermira, Inc. for study participation. Alexander Nast is an employee of Charité – Universitätsmedizin Berlin, which received compensation from Dermira, Inc. for study participation; he was an advisory board member for Boehringer Ingelheim, carried out educational activities for Bayer and Novartis, and received research grants from Eli Lilly and Company, Pfizer, GSK, Plc., and MEDA. Lawrence Green is an investigator for Brickell Biotech, Inc. and an advisory board member and investigator for Dermira, Inc. Richard D. Mamelok is a consultant for Dermira, Inc. John Quiring is an employee of QST Consultations. Janice Drew is an employee of Dermira, Inc. David M. Pariser is a consultant and investigator for Brickell Biotech, Inc., Celgene Corporation, Dermira, Inc., LEO Pharma US, Novartis Pharmaceuticals, Promius Pharmaceuticals, Regeneron, and Valeant Pharmaceuticals International, a consultant for ATACAMA, Biofrontera AG, DUSA Pharmaceuticals, Inc., Sonofi, TDM SurgiTech, Inc., and TheraVida, and an investigator for Abbott Laboratories, Amgen, Asana Biosciences, Dermavant Sciences, Eli Lilly and Company, Merck & Co., Inc., Novo Nordisk A/S, Ortho Dermatologics, Peplin Inc., Pfizer Inc., Photocure ASA, and Stiefel (as GSK company).

37. Paik J. Sofpironium Bromide: First Approval. Drugs. 2020;80(18):1981–6. Epub 2020/11/26. doi: 10.1007/s40265-020-01438-1. PubMed PMID: 33236266.

38. Kontochristopoulos G, Markantoni V, Agiasofitou E, Platsidaki E, Kouris A, Campanati A, et al. Treatment of primary axillary hyperhidrosis with a cream formulation of oxybutynin chloride 10. Journal of the European Academy of Dermatology and Venereology : JEADV. 2021;35(8):e524–e6. doi: 10.1111/jdv.17297. PubMed PMID: 33893675.

39. Chabicovsky M, Winkler S, Soeberdt M, Kilic A, Masur C, Abels C. Pharmacology, toxicology and clinical safety of glycopyrrolate. Toxicol Appl Pharmacol. 2019;370:154–69. Epub 2019/03/25. doi: 10.1016/j.taap.2019.03.016. PubMed PMID: 30905688.

40. Abels C, Soeberdt M, Kilic A, Reich H, Knie U, Jourdan C, et al. A glycopyrronium bromide 1% cream for topical treatment of primary axillary hyperhidrosis: efficacy and safety results from a phase IIIa randomized controlled trial. The British journal of dermatology. 2021;185(2):315–22. doi: 10.1111/bjd.19810. PubMed PMID: 33445205.

41. Nasir A, Bissonnette R, Maari C, DuBois J, Pene Dumitrescu T, Haddad J, et al. A phase 2a randomized controlled study to evaluate the pharmacokinetic, safety, tolerability and clinical effect of topically applied Umeclidinium in subjects with primary axillary hyperhidrosis. J Eur Acad Dermatol Venereol. 2018;32(1):145–51. Epub 2017/10/22. doi: 10.1111/jdv.14651. PubMed PMID: 29055158.

42. Shamseer L, Moher D, Clarke M, Ghersi D, Liberati A, Petticrew M, et al. Preferred reporting items for systematic review and meta-analysis protocols (PRISMA-P) 2015: elaboration and explanation. BMJ (Clinical research ed). 2015;350:g7647. doi: 10.1136/bmj.g7647. PubMed PMID: 25555855.

43. Liu HN WH, Yao C, Chen YZ, Liu TT. Advanced methods of data extraction for continuous outcomes in meta-analysis. Chinese Journal of Evidence-based Medicine. 2017;17(01):117–21.

44. Cumpston M, Li T, Page MJ, Chandler J, Welch VA, Higgins JP, et al. Updated guidance for trusted systematic reviews: a new edition of the Cochrane Handbook for Systematic Reviews of Interventions. The Cochrane database of systematic reviews. 2019;10:ED000142. doi: 10.1002/14651858.ED000142. PubMed PMID: 31643080.

45. Guyatt GH, Oxman AD, Vist GE, Kunz R, Falck-Ytter Y, Alonso-Coello P, et al. GRADE: an emerging consensus on rating quality of evidence and strength of recommendations. BMJ (Clinical research ed). 2008;336(7650):924–6. doi: 10.1136/bmj.39489.470347.AD. PubMed PMID: 18436948.

46. del Boz J. Systemic treatment of hyperhidrosis. Actas Dermosifiliogr. 2015;106(4):271–7. Epub 2015/02/02. doi: 10.1016/j.ad.2014.11.012. PubMed PMID: 25638324.

