## Supplementary material for "Topical anticholinergic medications for primary hyperhidrosis: A protocol for a systematic review and meta-analysis": S2 Appendix. The search strategy for PubMed

| | |
|---|---|
| #1 | "Hyperhidrosis"[Mesh] |
| #2 | "Cholinergic Antagonists"[Mesh] |
| #3 | (((((((((((((((((((((((((((((Antagonists, Cholinergic[Title/Abstract]) OR (Cholinergic Receptor Antagonist[Title/Abstract])) OR (Antagonist, Cholinergic Receptor[Title/Abstract])) OR (Receptor Antagonist, Cholinergic[Title/Abstract])) OR (Cholinergic-Blocking Agent[Title/Abstract])) OR (Agent, Cholinergic-Blocking[Title/Abstract])) OR (Cholinergic Blocking Agent[Title/Abstract])) OR (Cholinolytic[Title/Abstract])) OR (Cholinolytics[Title/Abstract])) OR (Acetylcholine Antagonist[Title/Abstract])) OR (Antagonist, Acetylcholine[Title/Abstract])) OR (Cholinergic Antagonist[Title/Abstract])) OR (Antagonist, Cholinergic[Title/Abstract])) OR (Cholinergic Receptor Antagonists[Title/Abstract])) OR (Antagonists, Cholinergic Receptor[Title/Abstract])) OR (Receptor Antagonists, Cholinergic[Title/Abstract])) OR (Cholinergic-Blocking Agents[Title/Abstract])) OR (Agents, Cholinergic-Blocking[Title/Abstract])) OR (Cholinergic Blocking Agents[Title/Abstract])) OR (Acetylcholine Antagonists[Title/Abstract])) OR (Antagonists, Acetylcholine[Title/Abstract])) OR (Anticholinergic Agents[Title/Abstract])) OR (Agents, Anticholinergic[Title/Abstract])) OR (Anticholinergic Agent[Title/Abstract])) OR (Agent, Anticholinergic[Title/Abstract])) OR (Anticholinergics[Title/Abstract])) OR (Anti-Cholinergics[Title/Abstract])) OR (Anti Cholinergics[Title/Abstract])) OR (Anti-Cholinergic[Title/Abstract])) OR (Anticholinergic[Title/Abstract]) |
| #4 | "glycopyrronium tosylate" [Supplementary Concept] |
| #5 | (3-(2-Cyclopentyl(hydroxy[Title/Abstract])phenylacetoxy)-1,1-dimethylpyrrolidinium 4-methylbenzenesulfonate hydrate (1:1:1)) OR (Qbrexza[Title/Abstract]) OR (Topical oxybutinin [Title/Abstract]) |
| #6 | #2 OR #3 OR #4 OR #5 |
| #7 | "Administration, Topical"[Mesh] |
| #8 | ((((((((Drug Administration, Topical[Title/Abstract]) OR (Topical Drug Administration[Title/Abstract])) OR (Topical Administration[Title/Abstract])) OR (Administrations, Topical[Title/Abstract])) OR (Topical Administrations[Title/Abstract])) OR (Administration, Topical Drug[Title/Abstract])) OR (Administrations, Topical Drug[Title/Abstract])) OR (Drug Administrations, Topical[Title/Abstract])) OR (Topical Drug Administrations[Title/Abstract]) |
| #9 | #7 OR #8 |
| #10 | "randomized controlled trial"[Publication Type] OR "controlled clinical trial"[Publication Type] OR "randomized"[Title/Abstract] OR "randomized"[Title/Abstract] OR "placebo"[Title/Abstract] OR "drug |

| | |
|---|---|
| | therapy"[MeSH Subheading] OR "randomly"[Title/Abstract] OR "trial"[Title/Abstract] OR "groups"[Title/Abstract] |
| #11 | #1 AND #6 AND #9 AND #10 |
